# Prevalence of SARS-CoV-2 Antibodies Among Healthcare Workers at a Tertiary Academic Hospital in New York City

**DOI:** 10.1101/2020.05.27.20090811

**Authors:** Mayce Mansour, Emily Leven, Kimberly Muellers, Kimberly Stone, Damodara Rao Mendu, Ania Wajnberg

## Abstract

**Background:** SARS-CoV-2 antibody testing is important for understanding immunity prevalence, and may have implications for healthcare workers (HCW) during the SARS-CoV-2 pandemic.

**Methods:** We conducted immunologic testing of healthcare workers to determine the prevalence of SARS-CoV-2 IgG in this population. HCW were advised to wait at least two weeks from time of symptom onset or suspected exposure before undergoing testing. All participants were self-reported asymptomatic for at least three days at the time of testing.

**Results:** Two hundred eighty-five samples were collected from March 24, 2020 to April 4, 2020. The average age of participants was 38 years (range 18-84), and 54% were male. Thirty-three percept tested IgG positive, 3% tested weakly positive, and 64% tested negative. Neither age nor sex was associated with antibody development.

**Conclusion:** Thirty-six percent of HCW had IgG antibodies to SARS-CoV-2, reflecting the high exposure of inpatient and ambulatory frontline staff to this viral illness, most of whom had minimal symptoms and were working in the weeks preceding testing. While we continue to recommend standard protective precautions per CDC guidelines for all HCW, HCW with SARS-CoV-2 IgG may become our safest frontline providers as we learn if our IgG antibodies confer immunity. Knowing IgG antibody status may ease concerns regarding personal risk as this pandemic continues.

## Introduction

SARS-CoV-2 antibody testing is important for understanding immunity prevalence, and may have implications for healthcare workers (HCW) during the SARS-CoV-2 pandemic. While it remains unknown whether SARS-CoV-2 antibody formation confers immunity, based on patterns seen in other viral illnesses, it is possible IgG presence may protect against reinfection.

HCW in the Mount Sinai Health System, a large, urban, academic tertiary care center, are at high risk for disease exposure as they live in areas with high rates of community spread and work in environments predominantly caring for SARS-CoV-2 patients. (1) Given limited availability of PCR testing, many HCW self-diagnose illness and self-isolate until resolution of symptoms, in accordance with Department of Health guidelines. (2)

We conducted immunologic testing of HCW to determine the prevalence of SARS-CoV-2 IgG in this population.

## Methods

We collected serum IgG antibody titers from self-referred HCW throughout our health system using a serologic ELISA assay. (3) In week one, we tested HCW in departments with the greatest exposure to aerosolized SARS-CoV-2 (i.e., emergency medicine, critical care, anesthesiology), and in week two tested all interested HCW with direct patient exposure. HCW were advised to wait at least two weeks from time of symptom onset or suspected exposure before undergoing testing. (4) All participants were self-reported asymptomatic for at least three days at the time of testing. Serum IgG titers were considered “positive” if detected at dilutions of 1:320 or greater and “weakly positive” if detected at dilutions of 1:50 to 1:160. Titers of 1:320 or greater were eligible for serum plasma donation. One-way ANOVA test and Fisher’s exact test were used to compare results among groups. Specimens were collected as part of our convalescent plasma donor identification and treatment program. This study received IRB exemption.

## Results

Two hundred eighty-five samples were collected from March 24, 2020 to April 4, 2020. The average age of participants was 38 years (range 18-84), and 54% were male. Thirty-three percept tested IgG positive, 3% tested weakly positive, and 64% tested negative. Neither age nor sex was associated with antibody development (Table 1). Nine percent were Ab positive in week one versus 44% in week two.

**Table 1.**

|  | **Health Care Worker Antibody Results** |  |  |  | **p-value** |
| --- | --- | --- | --- | --- | --- |
|  | **All (N=285)** | **Ab+ (N=93)** | **Ab weak+ (N=9)** | **Ab- (N=183)** |  |
| Age, Mean (SD) | 38.36 (10.81) | 37.15 (12.89) | 42.67 (8.05) | 38.63 (9.96) | 0.291 |
| Age, Range | 18 - 84 | 18 - 84 | 31 - 55 | 23 - 66 |  |
| Gender |  |  |  |  | 0.168 |
| Male, N (valid %) | 111 (54) | 53 (62) | 4 (50) | 54 (48) |  |
| Female, N (valid %) | 95 (46) | 33 (38) | 4 (50) | 58 (52) |  |
| Test Results, N (%) |  |  |  |  |  |
| Ab- | 183 (64) | - | - | - |  |
| Ab weak+ | 9 (3) | - | - | - |  |
| Ab+ | 93 (33) | - | - | - |  |

## Discussion

Thirty-six percent of HCW had IgG antibodies to SARS-CoV-2, reflecting the high exposure of inpatient and ambulatory frontline staff to this viral illness, most of whom had minimal symptoms and were working in the weeks preceding testing. Interestingly, while HCW in the first week of our study were in high risk departments, a larger proportion of HCW in the second week tested positive, likely reflecting the longer time course required for antibody development and the rise in community incidence. A limitation of our study was HCW who were eager to learn their antibody status may have presented too soon after exposure, leading to potential false negative testing. Additionally, it is possible that HCW with higher suspicion of infection were more likely to self-refer, potentially overestimating the rate of antibody positivity.

While we continue to recommend standard protective precautions per CDC guidelines for all HCW, HCW with SARS-CoV-2 IgG may become our safest frontline providers as we learn if our IgG antibodies confer immunity. Knowing IgG antibody status may ease concerns regarding personal risk as this pandemic continues. The next step will be to screen a larger proportion of our workforce in order to better stratify prior exposure based on work environment. Additionally, serial antibody testing will help us better understand duration of IgG response.

## Data Availability

NA

## Acknowledgements

Thank you to the medical students who assisted with outreaching and following up with our participants.

**Figures: NA**

